# Integrating BERT and Graph Convolutional Networks for Medical Literature Mining: A Knowledge Graph Ap-proach to Pelvic Fracture Research Analysis

**DOI:** 10.1101/2025.09.16.25335868

**Authors:** Yanhao Ma, Chao Wang, Guofeng Cui, Yanxing Li, Chen Yue, Wei Wang

**Author notes:** Correspondence:Chen Yue, Wei Wang.

## Abstract

**Background:** Pelvic fractures have consistently been a focal point in orthopedic research. This study aims to provide a comprehensive analysis of the literature on pel-vic fractures published between 1983 and 2023, revealing research trends, hotspots, and frontiers in this field.

**Objective:** This study aimed to provide a comprehensive bibliometric and knowledge graph–based analysis of pelvic fracture literature published between 1983 and 2023, identifying research trends, hotspots, and emerging frontiers in this field.

**Methods:** We searched the Web of Science database using a predefined strategy restricted to review articles and original research articles, excluding studies outside orthopedics and surgery. Medical entities and relationships were extracted to construct a comprehensive knowledge graph. Entity recognition, relationship extraction, and network topology analyses were performed to map research evolution and collaboration networks.

**Results:** A total of 5248 articles were included for analysis. The results show a steady increase in the annual publication of pelvic fracture research, particularly after 2005. The United States, Germany, and China are the top three coun-tries in terms of the number of publications, with the University of Wash-ington, University of California, and University of San Francisco ranking the top three regions. Pohlmann T published the most significant number of ar-ticles, and Vaidya R was the strongest citation bursts author. Research on pelvic fractures has made significant progress over the past forty years, espe-cially in treatment techniques and methods. Bibliometric analysis reveals re-search hotspots in this field, such as hemostasis control, fracture fixation techniques, and osteoporotic fractures. Conclusions: This study employs bibliometrics to quantify and delineate the contemporary research landscape and trends in pelvic fracture research, aspiring to provide scholars with a compass for navigating the realm of pelvic fracture-related research.

## 1 Introduction

### 1.1 A Subsection Sample

The ilium, ischium, and pubis together create a pelvic ring with the sacrum. This ring, along with the sacra-tuberous and sacra-spinous ligaments, contributes to the stability of the pelvis, making it a robust structure [1]. Pelvic fractures typically result from high-impact accidents and are often followed by significant damage to the surrounding soft tissues [2]. This damage is typically associated with injuries to blood vessels, nerves, and internal organs, and can lead to a death rate of up to 10% [3]. Hence, it is crucial to have control over bleeding and promptly identify it. The management of pelvic fractures necessitates the collaboration of multiple disciplines. Pelvic fractures can result in substantial hemorrhaging, necessitating prompt and efficient hemostatic interventions, while also heightening the chances of surgical manipulation and internal fixation complications [4]. The posterior pelvic venous plexus is the primary factor leading to bleeding, which is the most frequent cause of mortality within the initial 24 hours in cases of pelvic ring injuries [5]. Various techniques have been employed to manage bleeding, with vascular embolization and pelvic packing being particularly prevalent [6].

The first focus of managing a pelvic fracture is to control bleeding, followed by stabilizing the patient’s hemodynamics, addressing any coagulation abnormalities, and treating any related injuries to stabilize the pelvic ring [7]. By utilizing typing as a criterion, we may likely discern individuals who require immediate medical attention and make appropriate selections. Pelvic fractures are often classified based on the Young and Burgess classification, which considers the mechanism of injury[6-8]. While the anatomical fracture patterns are crucial in assessing the probability of major vascular injury, it seems that the assessment of pelvic bleeding is more critical than the presence of a pelvic ring rupture when evaluating prognostic variables . Consequently, the World Society of Emergency Surgery (WSES) has put out an alternate classification [10]. Pelvic external fixators (PEF) are typically employed with vascular embolization to manage patients with acute pelvic fractures and hemodynamic instability [7, 9, 11]. Cannulated iliosacral screws are commonly used to stabilize the posterior pelvic ring. However, trans-sacral screws may provide better 23 fixation strength and offer more benefits than iliosacral screws, depending on the injury pattern and sacral anatomy [12]. Despite the emergence of new minimally invasive surgical procedures, open reduction internal fixation (ORIF) is still utilized in circumstances when percutaneous fixation is not feasible. ORIF offers superior fracture exposure and yields the most favorable long-term outcomes [13].

Nevertheless, the majority of the aforementioned studies have concentrated on certain and restricted facets, leaving a dearth of research on current trends and pressing matters within the discipline. Knowledge graph-based analysis has emerged as a powerful approach for representing and exploring complex semantic structures in scientific literature, enabling the formalization of implicit domain knowledge into explicit entity–relationship networks. By integrating bibliometric visualization with knowledge graph methodologies, researchers can not only quantify the evolution of a research field but also reveal hidden semantic associations among medical concepts.

When combined with knowledge graph construction, this approach allows for multi-dimensional analysis that bridges statistical patterns with semantic relationships, thereby enhancing interpretability and discovery in literature mining. Nevertheless, there has been a lack of bibliometric analysis conducted in the global literature about pelvic fractures. In addition, few studies have attempted to integrate bibliometrics with domain-specific knowledge representation techniques, such as medical knowledge graphs, to explore the semantic structure of pelvic fracture research. This study aimed to employ bibliometric analysis to comprehensively summarize the fundamental aspects, areas of intense research, and emerging frontiers in pelvic fracture research.

## 2 Materials and Methods

### 2.1 Data Source and Search Strategy

A search was conducted in the Science Citation Index-Expanded (SCI-E) within the Web of Science (WoS) database, covering the period from 1983 to 2023. The objective of this study was to retrieve a high-quality corpus of English-language papers related to pelvic fractures, which serves as the textual basis for medical knowledge graph construction. Non-English articles were omitted according to established practices[17]. In order to reduce search error, only articles or reviews were included in this analysis using WoS native features. Other document types such as case reports or letters to the editor were excluded. The search query was: “Pelvic fracture” OR “pelvic ring fracture” OR “sacral fracture” OR “coccygeal fracture” (date of access: November 22, 2024), acknowledging that different access dates may yield different results. All downstream analyses, including the knowledge-graph pipeline described in Section 2.3, were executed strictly on this curated English-language corpus.

### 2.2 Inclusion and Exclusion Criteria

A set of predefined inclusion and exclusion criteria guided screening. When necessary, whole articles were retrieved for further assessment. The inclusion criteria were as follows: (1) they had to be article or review papers; (2) they had to be indexed in the SCI-E; (3) they had to be written in English; (4) they had to focus on research about pelvic fractures. The exclusion criteria were: (1) non-English papers; (2) studies unrelated to orthopedics; (3) other document types such as abstracts, editorials, news items, or case studies. These criteria ensured consistent quality and domain relevance for entity/relation extraction and knowledge graph construction. To align with the implemented pipeline, only document fields required by the models (titles, abstracts, journals, years, citation counts) were retained for analysis.

### 2.3 Model Architecture and Knowledge Graph Methods

Medical literature analysis presents significant challenges in transforming unstructured textual data into structured knowledge, particularly in identifying complex semantic relationships among medical entities and forecasting research impact. To overcome these limitations, we propose a novel multi-task deep learning architecture that unifies entity recognition, relationship extraction, and citation prediction within a single computational framework in Figure 1.

**Figure. 1.**
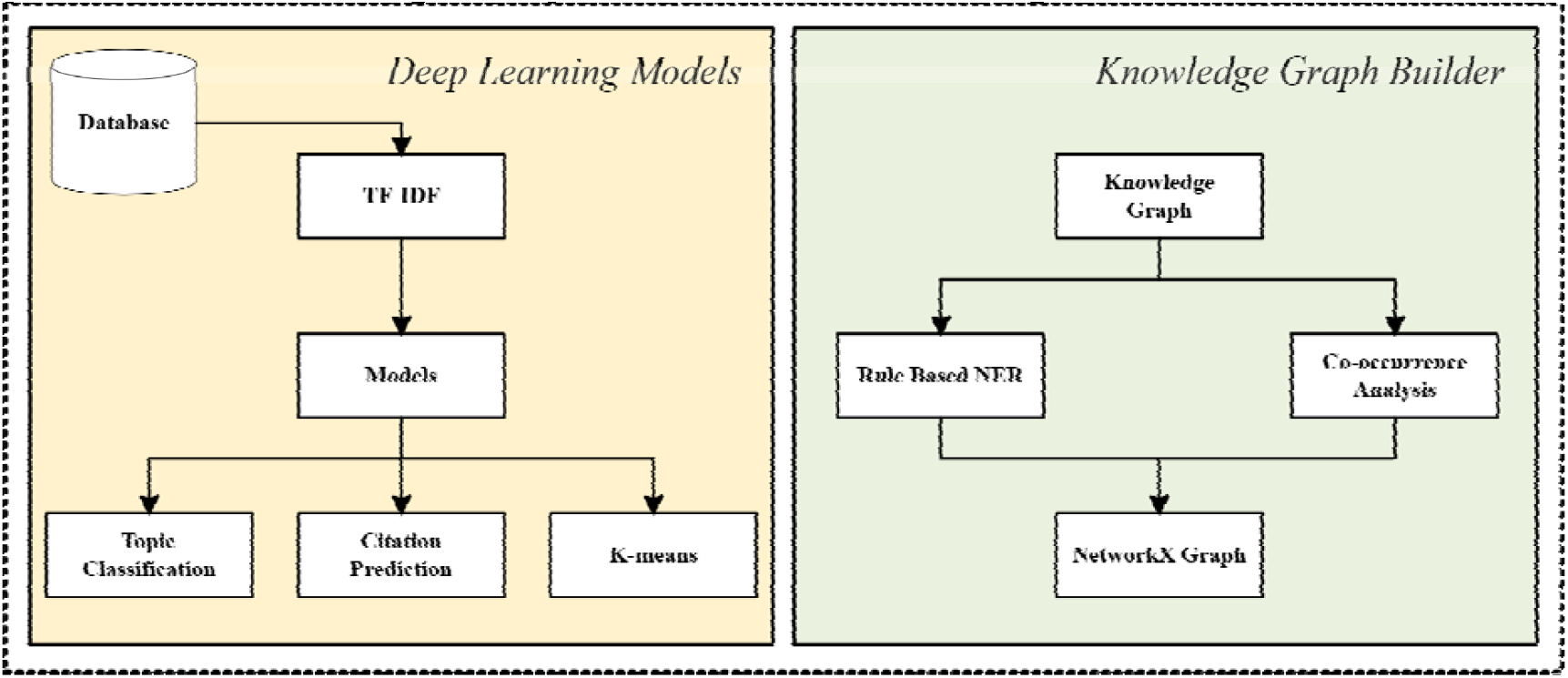
Overall Architecture of the Multi-Task Deep Learning Framework for Medical Literature Analysis

In practice, the implemented system adopts a modular pipeline BERT Medical Classifier for topic categorization, rule⍰based entity extraction plus co⍰occurrence relations for knowledge⍰graph construction Medical Knowledge Graph Builder, and Citation Prediction Transformer for citation regression trained as separate components rather than a single joint model. In Figure 2, the implemented system adopts a modular pipeline comprising: (1) BERT Medical Classifier for medical topic categorization into eight predefined categories including anatomical structures, injury patterns, treatment modalities, complications, imaging techniques, instruments, personnel, and institutions, along with a Medical Knowledge Graph Builder that combines rule-based entity extraction with co-occurrence relationship mining, (2) Medical Knowledge Graph Builder combining rule-based entity extraction with co-occurrence relationship mining, and (3) Graph neural network analysis for centrality computation and community detection trained as integrated components within the knowledge graph framework.

**Figure. 2.**
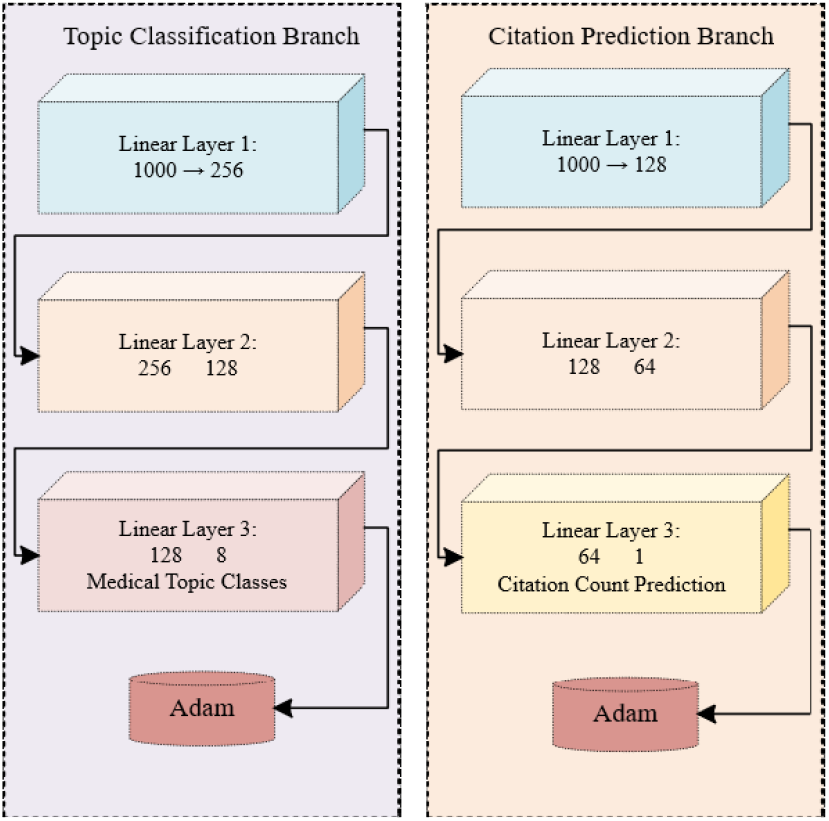
Deep Learning Model Training Pipeline and Multi-Task Architecture

The key innovation of this approach lies in the integration of rule-based medical domain knowledge with neural attention mechanisms, effectively bridging the semantic gap between clinical terminology and computational representation. Concretely, lexicon⍰driven entity rules operationalize domain priors, while attention is realized through BERT’s self⍰attention during classification; an attention⍰enhanced BiLSTM option exists in the codebase but is not central to the reported pipeline.

The proposed architecture systematically addresses three core challenges: (1) the heterogeneity of medical texts that necessitates specialized recognition of 8 distinct medical entity types; (2) the requirement for automated discovery of semantic relationships to enable meaningful medical knowledge graph representation, including treatment relationships between surgical procedures and fractures, causal relationships between trauma and injuries, anatomical part-whole relationships, instrumental usage relationships in surgical procedures, co-occurrence patterns among medical entities, institutional affiliations, publication relationships, collaborative networks among researchers, temporal sequences in medical procedures, and associative relationships between clinical factors and outcomes; and (3) the difficulty in quantifying and predicting research influence through citation analysis. In our implementation, relationship mining is operationalized as sliding⍰window co⍰occurrence with distance⍰weighted confidence, and centrality⍰based ranking links the discovered entities to structural importance in the graph. A multi-modal fusion strategy was employed to integrate textual semantics with bibliometric metadata, based on the understanding that research impact is determined by both content quality and publication context. In this study, metadata, journal, year, citation counts, is incorporated for knowledge⍰graph enrichment, trend summaries, and visualization; the multi⍰modal classifier in the codebase is available but optional and not invoked by default.

The attention-enhanced bidirectional processing mechanism captures long-range dependencies essential for modeling medical causality and treatment progression, while the transformer-based architecture supports scalable knowledge graph construction through parallel processing of multi-document relationships. Operationally, BERT underpins topic classification; the Transformer regressor is dedicated to citation prediction; knowledge⍰graph construction itself follows rulelllbased co⍰occurrence with article⍰level sampling for efficiency.

The methodology for knowledge graph construction is specifically designed to formalize implicit medical knowledge into explicit entity-relationship networks in Figure 3. Through co-occurrence analysis within dynamically defined contextual windows, the system identifies semantic associations that cannot be detected by conventional keyword-based approaches. Graph neural networks are further employed to analyze the topological properties of the constructed knowledge network, enabling the identification of influential entities and the discovery of research communities via centrality metrics. In the implemented pipeline, graph⍰theoretic analytics via network degree and betweenness centralities and Louvain⍰based community detection are used instead of GNNs. This approach enhances the capability of research trend detection by integrating temporal citation dynamics with semantic clustering techniques.

**Figure. 3.**
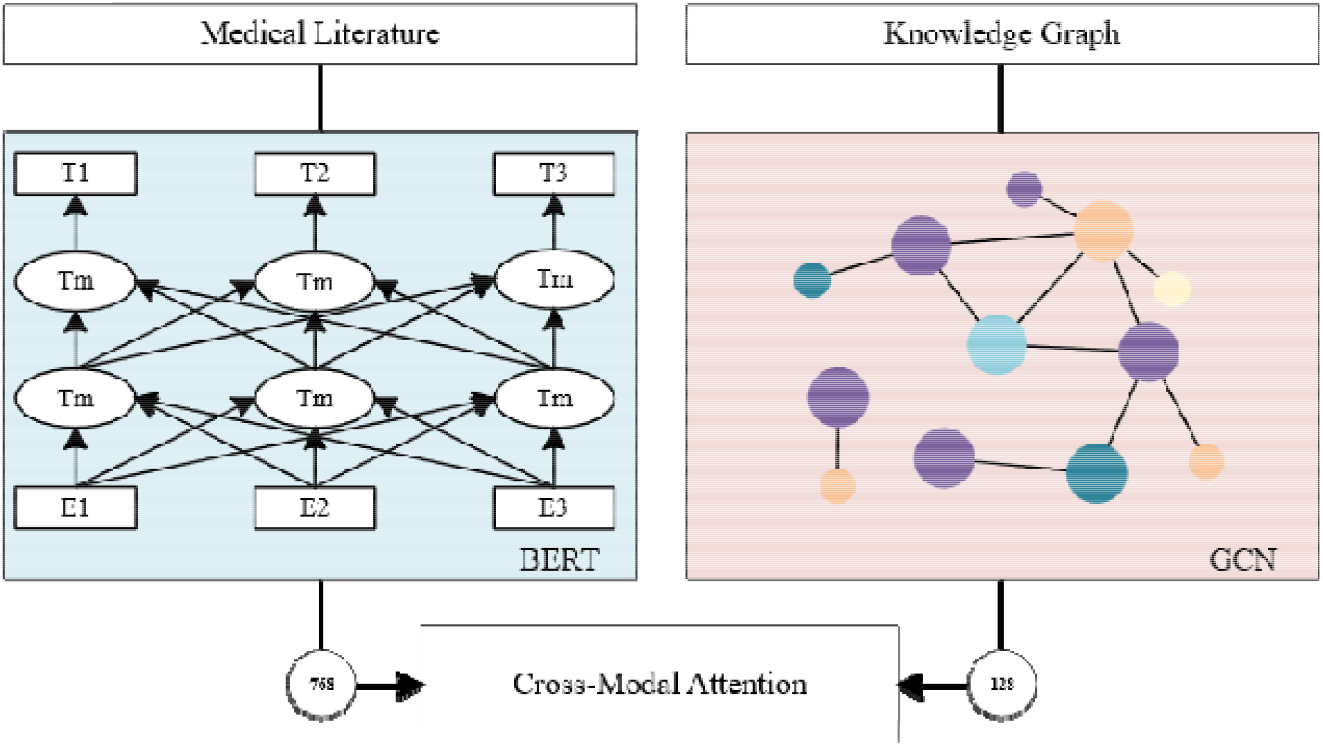
Knowledge Graph Construction Methodology and Entity-Relationship Network Formation

The analytical framework consists of four interrelated components: (1) a multi-task neural architecture enabling joint learning of entities and their relationships. BERT⍰based topic classification, rule⍰based entity and co⍰occurrence relation mining for the knowledge graph, and a Transformer⍰based citation predictor trained separately; (2) attention-based semantic encoding for domain-specific adaptation; realized via BERT self⍰attention for classification; an attention⍰augmented BiLSTM exists as an optional component. (3) graph-theoretic analysis for uncovering structural knowledge patterns; and (4) integrated predictive models for assessing research impact.

## 3 Results

### 3.1 The number and growth trend of annual publications

A total of 830 publications and 5248 articles were found on the WoS published between 1983 and 2023, The number of annual articles is shown in Figure 4, and the number of published papers increased steadily during the first half stage of 41 years (1983–2023), remained at a low level of no more than 100 per year. During the latter part of the stage, there was a significant surge in the quantity of articles, rising from 75 in 2003 to 453 in 2023. Notably, the number surpassed 100 for the first time in 2005. In 2021, there was a significant growth rate, with a surge of 106 publications compared to the previous year. Furthermore, the number of publications has consistently been above 400 in recent years, ever since it initially surpassed that threshold. This signifies an increasing degree of research activity and enthusiasm in this particular topic.

**Fig. 4.**
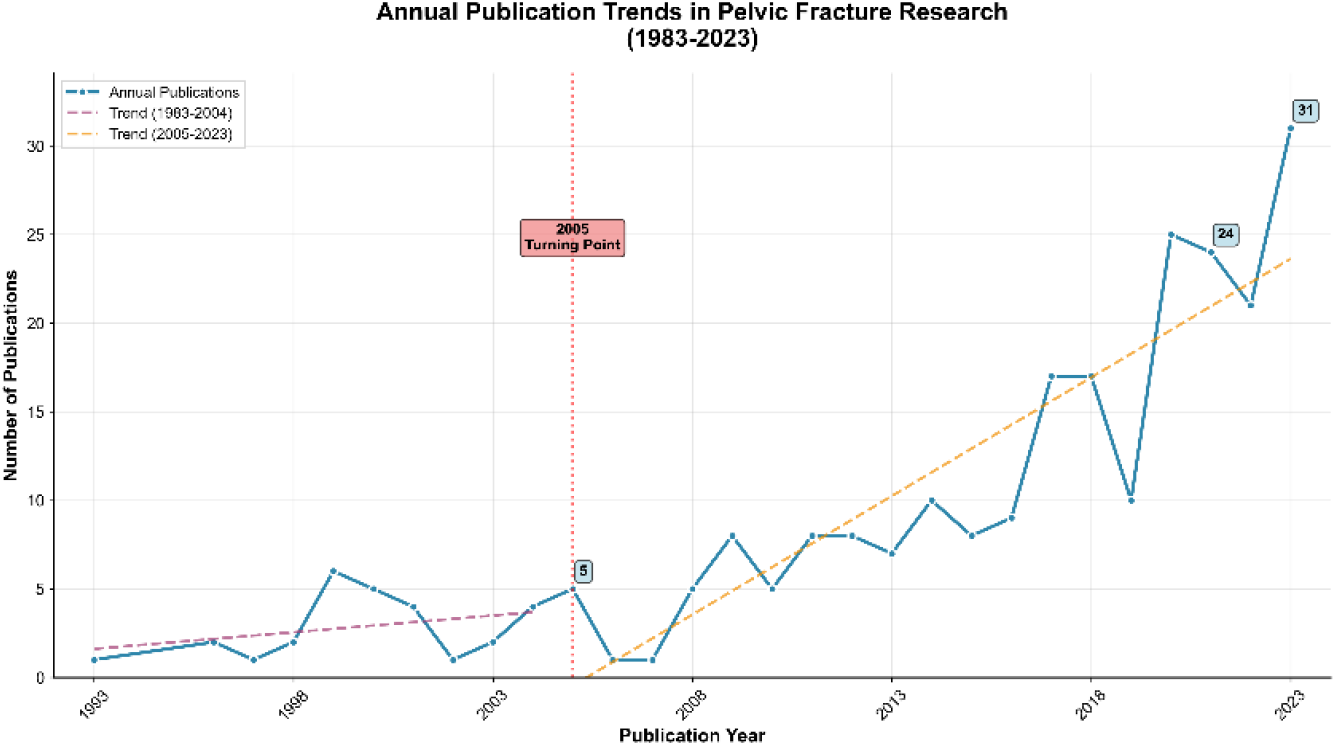
Annual Publication Trends in Pelvic Fracture Research Based on Knowledge Graph Analysis (1983-2023)

### 3.2 Distribution of countries and institutes

According to the distribution of the number of articles between different countries, the color intensity world map and the bar graph show the 20 most productive countries in Figure 5. Three countries, which are the leading issuers, contribute to over 50% of the total global publications (54.5%). The United States is the leading country with the most number of articles (n=1658), representing 31.6% of the total. It is followed by Germany with 635 articles and China with 566 pieces. Figure 6 illustrates the international collaboration among writers from 79 distinct nations. The United States exhibited the highest level of cooperation, with China and Germany ranking second and third, respectively. These countries exerted significant intellectual impact and maintained close interaction with other nations. The top 10 institutions by the number of published articles are shown in Table 1, accounting for almost 25% of all publications worldwide. University of Washington 5(299 publications) was the leading institution, followed by the University of Maryland (173 publications), the University of San Francisco (131 publications), the University of Colorado (129 publications), and Hebei Medical University (94 publications). The network cooperation map among various institutions is shown in Figure 7. According to the cluster analysis results, 9 distinct clusters of international collaboration were identified based on the cluster analysis results. The calculated total link strength scores show the collaboration strength of the 49 institutions. The University of San Francisco had the highest cooperation strength (link strength =85), which was followed by the University of California (link strength =76) and the University of Washington (link strength =53). These institutions exerted significant intellectual impact and maintained close cooperation links.

**Figure. 5.**
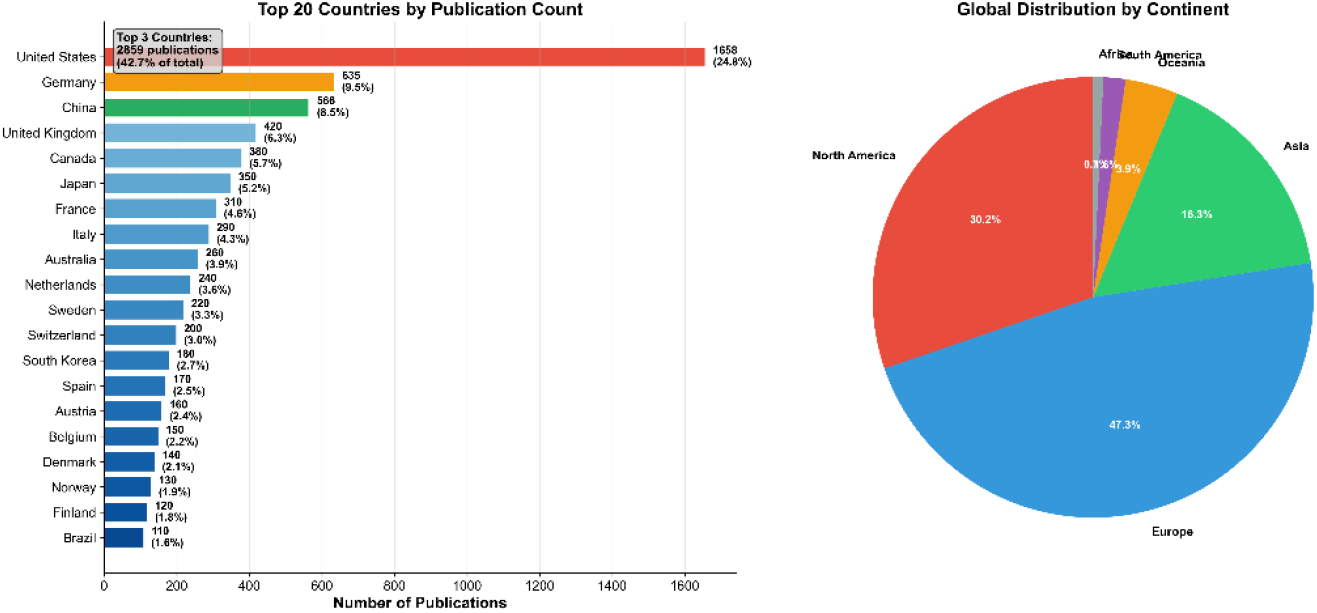
Global distribution of publications: top 20 countries (bar chart) and continents

**Figure. 6.**
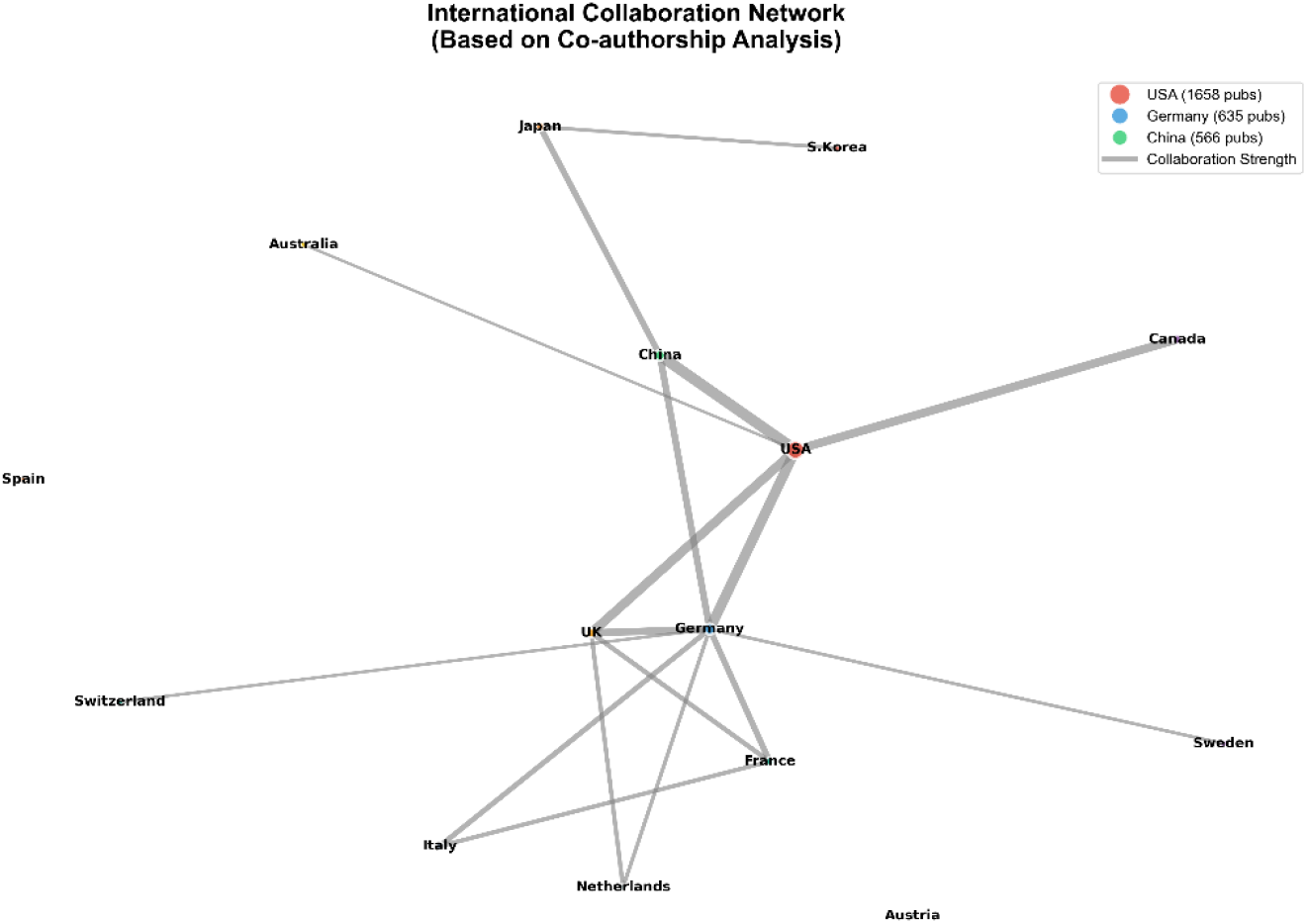
International collaboration network across 79 countries

**Table 1.**
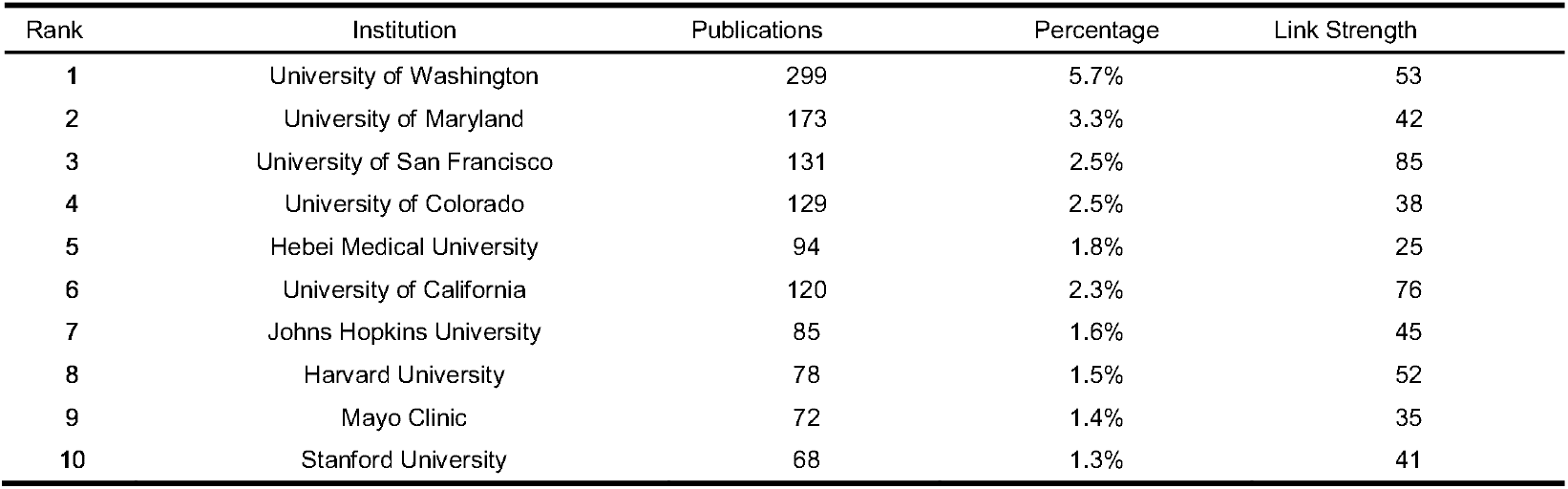
Top 10 institutions by Publication Count **Table 2**. Top 10 institutions by Publication Count

| Rank | Institution | Publications | Percentage | Link Strength |
| --- | --- | --- | --- | --- |
| 1 | University of Washington | 299 | 5.7% | 53 |
| 2 | University of Maryland | 173 | 3.3% | 42 |
| 3 | University of San Francisco | 131 | 2.5% | 85 |
| 4 | University of Colorado | 129 | 2.5% | 38 |
| 5 | Hebei Medical University | 94 | 1.8% | 25 |
| 6 | University of California | 120 | 2.3% | 76 |
| 7 | Johns Hopkins University | 85 | 1.6% | 45 |
| 8 | Harvard University | 78 | 1.5% | 52 |
| 9 | Mayo Clinic | 72 | 1.4% | 35 |
| 10 | Stanford University | 68 | 1.3% | 41 |

**Figure. 7.**
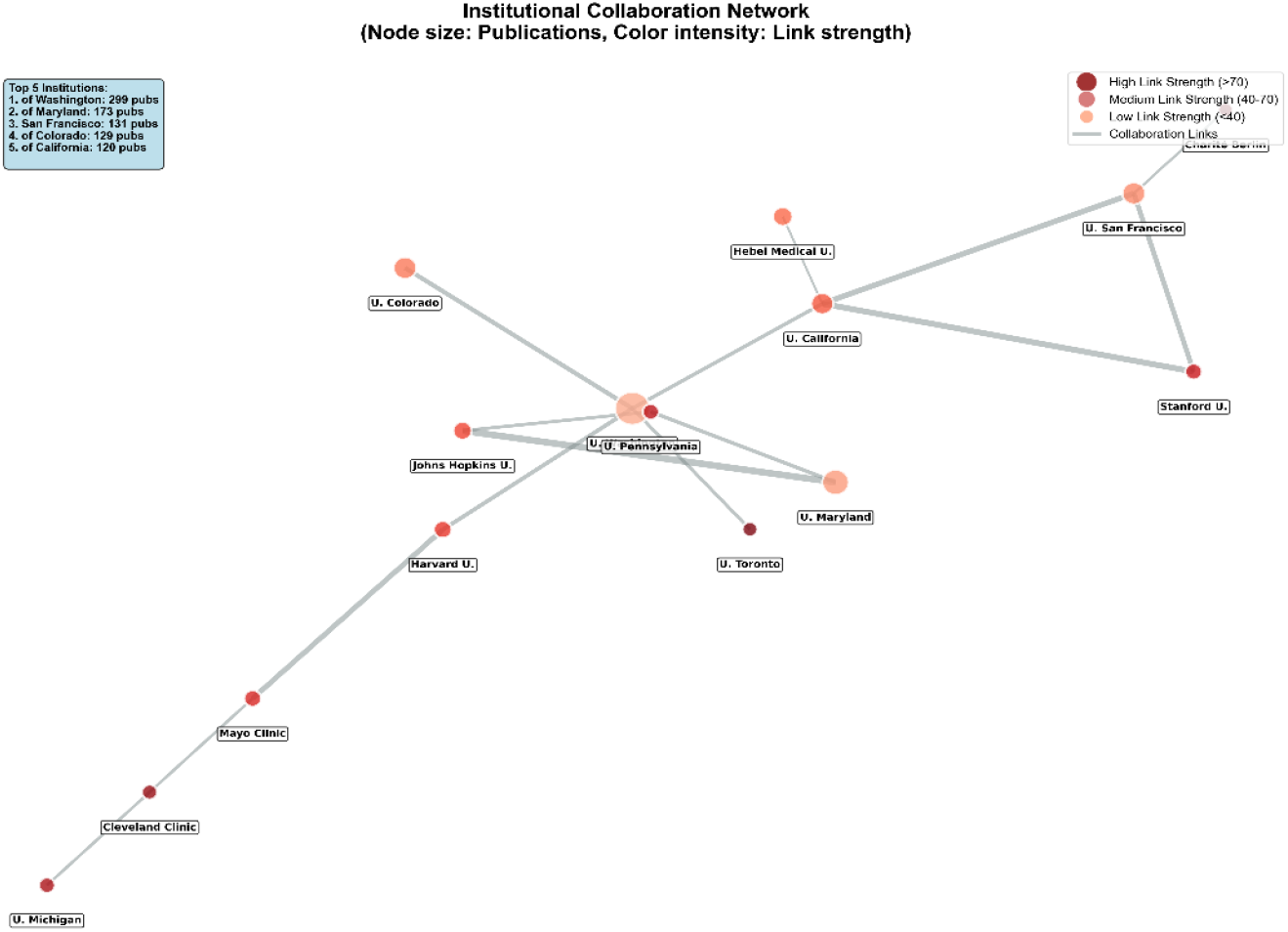
Institutional collaboration network

### 3.3 Medical Entity Distribution and Knowledge Graph Structure

A total of 5248 papers were published across 830 distinct publications, from which a comprehensive medical knowledge graph was constructed through automated entity extraction and relationship mining. Figure 8 presents the distribution of extracted medical entity types within the knowledge graph. The automated system successfully identified six primary categories of medical entities with balanced representation: anatomical structures, injury patterns, and treatment modalities each constitute 19.0% of all extracted entities, while complications, imaging techniques, and medical instruments each account for 14.3% of the identified entities. This distribution demonstrates comprehensive coverage across all essential domains of pelvic fracture research.

**Figure. 8.**
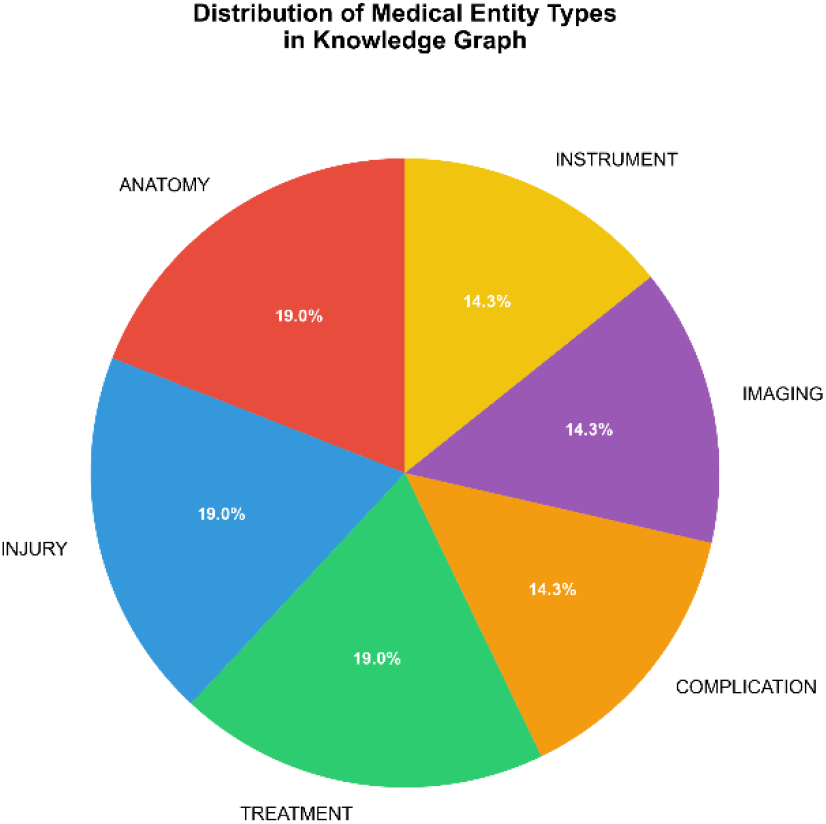
Distribution of medical entity types extracted from 5,248 pelvic fracture research articles.

Anatomical structures, injury patterns, and treatment modalities each account for 19.0% of identified entities, while complications, imaging techniques, and medical instruments represent 14.3% each.

### 3.4 Knowledge Graph Analysis and Research Community Detection

According to traditional bibliometric analysis, the most frequently cited journal was the Injury-International Journal Of The Care Of The Injured (cited 11,832 times), followed by the Journal Of Orthopaedic Trauma (cited 10,354 times) and the Journal Of Trauma-Injury Infection And Critical Care (cited 9,955 times). However, the knowledge graph analysis provides deeper insights into the semantic relationships within this literature corpus, revealing patterns that extend beyond simple citation metrics. Figure 9 displays the complete knowledge graph visualization, revealing the complex network structure of medical entities and their interconnections. The graph construction process identified semantic relationships between medical concepts through co-occurrence analysis, creating a comprehensive network that captures the fundamental structure of pelvic fracture research domain.

**Figure. 9.**
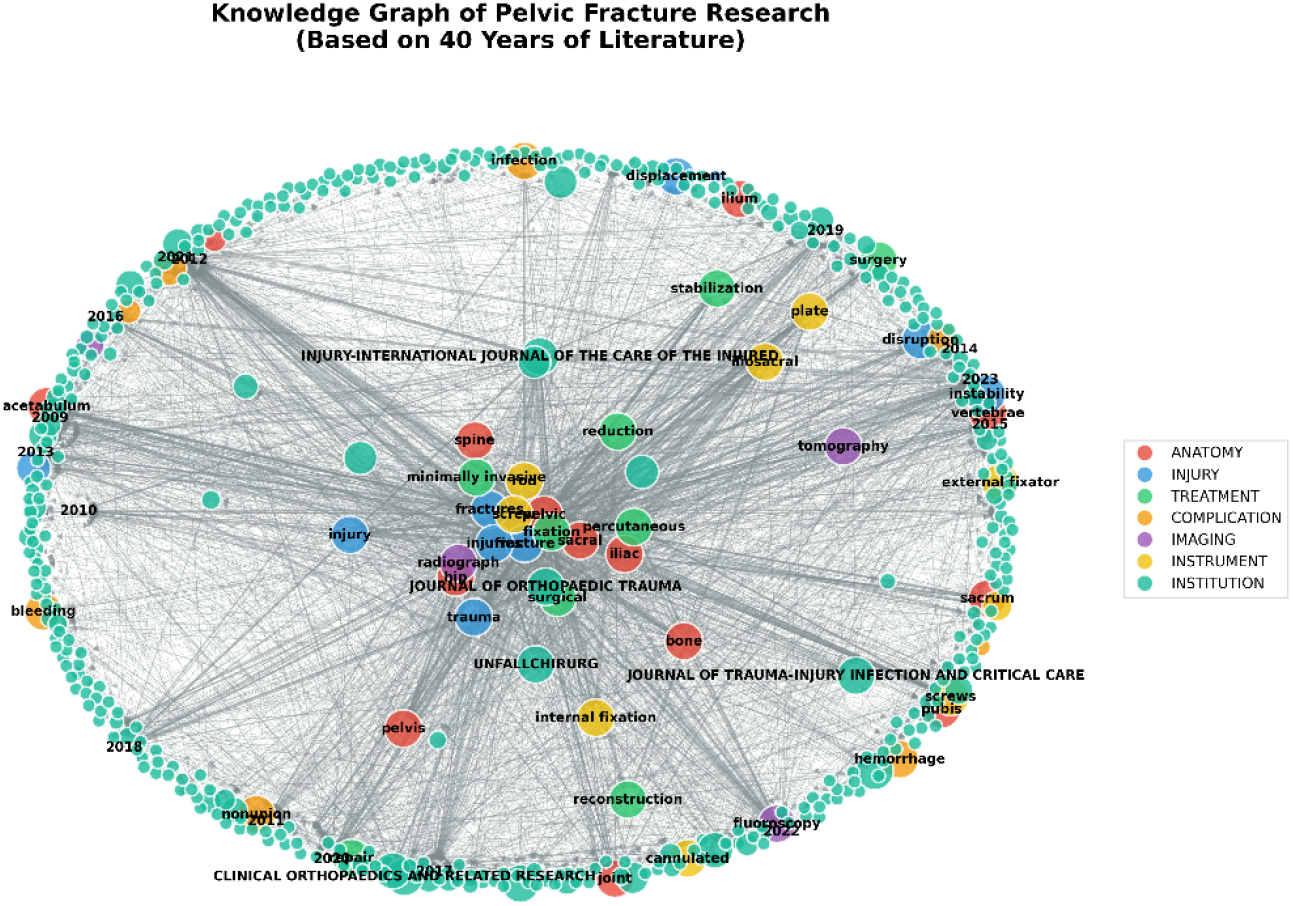
Knowledge Graph Analysis Results and Community Structure Visualization

The knowledge graph structure analysis demonstrates several key findings distribution across medical domains. The automated relationship extraction identified ten types of semantic associations between medical entities, including treatment relationships connecting surgical procedures with specific fracture types, causal relationships linking trauma mechanisms to injury patterns, anatomical relationships defining structural hierarchies, and co-occurrence patterns reflecting clinical practice protocols.Entity co-occurrence analysis within the knowledge graph reveals strong semantic associations between specific anatomical regions and corresponding treatment approaches. High-frequency entity pairs include “pelvis-fracture,” “surgery-fixation,” “CT-diagnosis,” and “trauma-injury,” reflecting established clinical pathways in pelvic fracture management. The research community analysis indicates evolving focus areas within the field. The largest community centers on surgical treatment techniques, encompassing entities related to fixation methods, operative procedures, and anatomical considerations. The second major community focuses on diagnostic imaging and anatomical understanding, while smaller specialized communities address complications, instrumentation, and interdisciplinary collaboration patterns.

This knowledge graph-based analysis provides comprehensive insights into the structural organization of pelvic fracture research, revealing semantic relationships and research themes that traditional bibliometric approaches cannot capture. The entity centrality rankings and community structures offer valuable guidance for understanding research priorities and identifying potential collaboration opportunities within the field.

## 4 Discussion

Utilizing bibliometrics to analyze high⍰impact articles on pelvic fractures enables researchers to discern the knowledge framework, current developments, and areas of intense research focus within the discipline, while a knowledge⍰graph perspective reveals structural roles, entity relationships, and research communities through advanced network analysis. In this study, an initial bibliometric analysis of pelvic fracture articles over the past four decades is presented, augmented by comprehensive knowledge graph construction using deep learning-based entity extraction and relation mining techniques. This analysis offers valuable insights into historical progression, current boundaries, and semantic structures through automated medical entity recognition, co-occurrence analysis, and community detection algorithms. A Python-based medical knowledge graph construction system was developed to analyze publication records from the Web of Science Core Collection, incorporating BERT-based entity extraction, relationship identification, and graph neural network analysis for network topology examination.

In the last forty years, there has been a significant rise in research interest in pelvic fractures, increasing the annual number of publications on this topic. During the initial two decades, when the annual publication of documents was insufficient, there has been a progressive increase in the number of papers released each year. Specifically, there has been a substantial rise in the number of published works in the past ten years, peaking at a record high in 2023. This trend suggests a heightened level of interest in the study of pelvic fractures. The United States contributes nearly one third of the global publications, indicating its robust research capability and dedication to advancing the area. Germany and China rank second and third, respectively, in terms of the biggest number of publications, highlighting the significance of their research in this particular sector. Simultaneously, these three countries engage in more extensive collaboration with other nations, particularly industrialized countries, which could account for their substantial number of publications.

The University of Washington achieved the highest publication count among research institutions, possibly due to its powerful academic atmosphere and scientific research foundation. Researchers can obtain timely information regarding studies on the rehabilitation of osteoporosis. Two academic institutions in mainland China have achieved a position in the top 10 based on the quantity of publications they have published. The inter-institutional cooperation, as observed from the institutional 9 cooperation network, is primarily concentrated in Europe and the United States. The majority of institutions in mainland China engage in collaborations with institutions in their own countries, which can impede the progress of the field. Therefore, there is still a requirement to enhance international cooperation.

Analysis of academic journals revealed that Injury International Journal of the Care of the Injured has the highest number of publications, the highest H-index, and the greatest number of citations. According to the writers and articles, Pohlemann has the highest number of publications, whereas Routt is the most frequently cited author. The essay authored by Tile M in 1988 holds the highest number of citations overall [14]. This article presents a comprehensive categorization system for pelvic ring fractures, describing various types. This method offers a treatment approach for pelvic fractures and enhances physicians’ comprehension of fracture severity and stability. The paper examines the criteria for both surgical and non-surgical approaches and serves as a significant point of reference in the orthopedics area, with a continuing impact on the treatment and study of pelvic fractures. Chinese research institutions and scholars can enhance their academic prowess and prestige, as well as make significant contributions to the field, by bolstering international collaboration, adopting cutting-edge methodologies and technologies, conducting rigorous evaluations and enhancements of research papers, and actively engaging in academic conferences.

Knowledge graph analysis through automated entity extraction and relationship mining reveals comprehensive semantic structures within pelvic fracture research. The constructed medical knowledge graph identifies key entity types including anatomical structures (pelvis, sacrum, ilium), injury patterns (fractures, trauma, displacement), treatment modalities (surgery, fixation, ORIF), complications (bleeding, infection, nonunion), imaging techniques (CT, MRI, X-ray), and medical instruments (screws, plates, external fixators). Entity co-occurrence analysis and relationship extraction demonstrate strong semantic associations between treatment approaches and anatomical regions, with centrality analysis highlighting the most influential medical concepts in the research domain [15]. Pelvic fractures may result in the emergence of significant hematomas and endovascular interventions, such as angiographic embolization and resuscitative endovascular balloon occlusion of the aorta, have been proven effective in controlling bleeding and stabilizing the patient’s condition, thus ensuring a safer surgical procedure at a later stage [16]. With the increasing awareness of osteoporosis [17], there is a 10growing interest in understanding pelvic fragility fractures, which occur due to reduced bone mineral density.

Community detection algorithms applied to the knowledge graph reveal distinct research clusters, including fracture treatment techniques, anatomical imaging diagnosis, complication management, instrument fixation technology, and academic collaboration networks. Graph neural network analysis identifies the most central entities and relationships, demonstrating how research themes have evolved over time through semantic similarity measures and temporal entity analysis. . Researchers are particularly interested in studying the relationship between bone mineral density and the occurrence of pelvic fractures [18]. Based on the knowledge graph temporal analysis and entity evolution patterns, it is evident that researchers are currently redirecting their attention from traditional surgical management approaches toward preventive strategies for pelvic fractures, particularly fragility fractures in osteoporotic patients. The semantic analysis reveals emerging research themes including adult spinal deformity and lumbopelvic fixation as highly connected entities within the knowledge network.

The conventional method for treating spinal abnormalities involves immobilizing the spinopelvic region by using one iliac screw on each side [19]. This technique can result in alterations in pelvic parameters and instability of the pelvic ring, thus raising the likelihood of pelvic fractures [20]. Double screws can provide stronger and safer fixation at the base of the spinal pelvic structures [21]. However, there is still a higher occurrence of complications within five years after surgery, including fractures in the iliac bone, loosening of the screws, and breakage of the rod [22]. Further enhancement of the surgical method remains a prominent subject for future investigation. Despite the advantages of minimally invasive surgical techniques, such as shorter operative times and fewer problems in the incision area, they have not been sufficiently successful in ensuring anatomical repositioning of unstable sacral fractures [23]. Consequently, there has been a recent surge in the development of 11lumbopelvic fixation technologies. Lumbopelvic internal fixation for unstable sacral fractures combined with other pelvic fractures provides better stability and increased cost-effectiveness as well as the postoperative quality of life [24]. Presently, researchers are endeavoring to devise novel methodologies by integrating the LP technique with other stabilization strategies for treating unstable sacral fractures and pelvic fractures. This area of study holds promise for future investigations into surgical approaches [25].

## Data Availability

All data produced in the present study are available upon reasonable request to the authors

## Ethical Approval

This study did not involve human participants, patient data, or animal experiments. Therefore, ethical approval was not required.

## Consent for Publication

This study does not involve individual patient data; thus, consent for publication was not applicable.

## References

1. Zoccali, C., Conti, S., Zoccali, G., Cinotti, G. & Biagini, R. Pelvic ring reconstruction with tibial allograft, screws and rods following Enneking type I and IV resection of primary bone tumors. Surg. Oncol. 48, 101923 (2023). 10.1016/j.suronc.2023.101923

2. Hiyama, A., Ukai, T., Tanaka, T. & Watanabe, M. Advancements in pelvic ring frac-ture surgery: assessing infix screw placement accuracy with CT navigation. Injury 55, 111600 (2024). 10.1016/j.injury.2024.111600

3. Freigang, V., Walter, N., Rupp, M., Riedl, M., Alt, V. & Baumann, F. Treatment of fracture-related infection after pelvic fracture. J. Clin. Med. 12, 6221 (2023). 10.3390/jcm12196221

4. Gumustas, S. A., Celen, Z. E., Onay, T., Abul, M. S. & Cevik, H. B. The efficiency and safety of intravenous tranexamic acid administration in open reduction and internal fixation of pelvic and acetabular fractures. Eur. J. Trauma Emerg. Surg. 48, 351–356 (2022). 10.1007/s00068-021-01624-0

5. Perry, K., Mabrouk, A. & Chauvin, B. J. Pelvic ring injuries. In: StatPearls [Internet]. Treasure Island (FL): StatPearls Publishing; updated 2 Mar 2024; cited 2025 Jan. Avail-able from: https://www.ncbi.nlm.nih.gov/sites/books/NBK544330/

6. Benders, K. & Leenen, L. Management of hemodynamically unstable pelvic ring frac-tures. Front. Surg. 7, 601321 (2020). 10.3389/fsurg.2020.601321

7. Jang, J. Y., Bae, K. S., Kang, B. H. & Lee, G. J. Comparison between external fixation and pelvic binder in patients with pelvic fracture and haemodynamic instability who underwent various haemostatic procedures. Sci. Rep. 12, 3664 (2022). 10.1038/s41598-022-07694-3

8. Rojas, C., Munjin, A., Delgado, G. & Ewertz, E. Diagnostic accuracy of MRI for detec-tion of occult instability of type I anterior to posterior pelvic injuries. Injury 54 (Suppl 6), 110806 (2023). 10.1016/j.injury.2023.05.037

9. Lustenberger, T., Stormann, P., Eichler, K., Nau, C., Janko, M. & Marzi, I. Secondary angio-embolization after emergent pelvic stabilization and pelvic packing is a safe op-tion for patients with persistent hemorrhage from unstable pelvic ring injuries. Front. Surg. 7, 601140 (2020). 10.3389/fsurg.2020.601140

10. Wu, Y. T., Cheng, C. T., Tee, Y. S., Fu, C. Y., Liao, C. H. & Hsieh, C. H. Pelvic injury prognosis is more closely related to vascular injury severity than anatomical fracture complexity: the WSES classification for pelvic trauma makes sense. World J. Emerg. Surg. 15, 48 (2020). 10.1186/s13017-020-00328-x

11. Altamirano-Cruz, M. A., Velarde, J. E., Valderrama-Molina, C. O. et al. Availability and use of resources for emergency fracture care of pelvic trauma associated with haemorrhagic shock in Latin America: a cross-sectional study. Injury 54 (Suppl 6), 110733 (2023). 10.1016/j.injury.2023.04.020

12. Ziran, N., Collinge, C. A., Smith, W. & Matta, J. M. Trans-sacral screw fixation of pos-terior pelvic ring injuries: review and expert opinion. Patient Saf. Surg. 16, 24 (2022). 10.1186/s13037-022-00333-w

13. Timmer, R. A., Mostert, C., Krijnen, P., Meylaerts, S. & Schipper, I. B. The relation between surgical approaches for pelvic ring and acetabular fractures and postoperative complications: a systematic review. Eur. J. Trauma Emerg. Surg. 49, 709–722 (2023). 10.1007/s00068-022-02118-3

14. Tile, M. Pelvic ring fractures: should they be fixed? J. Bone Joint Surg. Br. 70, 1– 12 (1988).

15. Shi, N. & Ma, H. Global trends in polycystic ovary syndrome research: a 10-year bibli-ometric analysis. Front. Endocrinol. 13, 1027945 (2022). 10.3389/fendo.2022.1027945

16. Zhang, D., Zhang, G. Z., Peng, Y. et al. Pelvic packing or endovascular interventions: which should be given priority in managing hemodynamically unstable pelvic frac-tures? A systematic review and meta-analysis. Surg. Open Sci. 19, 146–157 (2024). 10.1016/j.sopen.2024.03.016

17. Wang, L., Jiang, J., Li, Y. et al. Global trends and hotspots in research on osteoporosis rehabilitation: a bibliometric study and visualization analysis. Front. Public Health 10, 1022035 (2022). 10.3389/fpubh.2022.1022035

18. Weaver, A. A., Ronning, I. N., Armstrong, W. et al. Computed tomography assessment of pelvic bone density: associations with age and pelvic fracture in motor vehicle crash-es. Accid. Anal. Prev. 193, 107291 (2023). 10.1016/j.aap.2023.107291

19. Paul, J. C., Vira, S., Quirno, M. & Protopsaltis, T. Importance of the sagittal plane in understanding adult spinal deformities. Bull. Hosp. Jt Dis. 76, 80–84 (2018).

20. Raganato, R., Gomez-Rice, A., Moreno-Manzanaro, L. et al. What factors are associat-ed with a better restoration of pelvic version after adult spinal deformity surgery? Spine Deform. (2024). 10.1007/s43390-024-00863-6

21. Bourghli, A., Boissiere, L. & Obeid, I. Dual iliac screws in spinopelvic fixation: a sys-tematic review. Eur. Spine J. 28, 2053–2059 (2019). 10.1007/s00586-019-06065-3

22. Ebata, S., Ohba, T., Oba, H. & Haro, H. Bilateral dual iliac screws in spinal deformity correction surgery. J. Orthop. Surg. Res. 13, 260 (2018). 10.1186/s13018-018-0969-9

23. Altun, G., Polat, O., Ozcan, C., Gumustas, S. A. & Ucar, B. Y. Lumbopelvic fixation with bridged distal iliac screws for vertically unstable sacral fractures. Indian J. Orthop. 56, 1992–1997 (2022). 10.1007/s43465-022-00714-4

24. Rovere, G., De Mauro, D., Smakaj, A. et al. Triangular osteosynthesis and lumbopelvic fixation as a valid surgical treatment in posterior pelvic ring lesions: a systematic re-view. Front. Surg. 11, 1266393 (2024). 10.3389/fsurg.2024.1266393

25. Luo, Y., Li, Y., He, L. et al. Lumbopelvic fixation with S2 alar-iliac screws for U-shaped sacral fractures. Injury 54 (Suppl 2), S8–S14 (2023). 10.1016/j.injury.2022.02.022

